# Use of inhibitors of the renin angiotensin system and COVID-19 prognosis: a systematic review and meta-analysis

**DOI:** 10.1101/2020.05.19.20106799

**Authors:** Jessica Barochiner, Rocío Martínez

**Author notes:** **Corresponding author:** Dr. Jessica Barochiner. **Address:** Tte. Gral. Juan Domingo Perón 4190 (zip code: C1199ABB), Buenos Aires, Argentina. **Email:**.

## Abstract

**Background:** controversy has arisen in the scientific community on whether the use of renin angiotensin system (RAS) inhibitors in the context of COVID-19 would be of benefit or harmful. A meta-analysis of eligible studies comparing the occurrence of severe and fatal COVID-19 in infected patients who were under treatment with angiotensin converting enzyme inhibitors (ACEI) or angiotensin receptor blockers (ARB) vs no treatment or other antihypertensives was conducted.

**Methods:** PubMed, Google Scholar, the Cochrane Library, MedRxiv and BioRxiv were searched for relevant studies. Fixed-effect models or random-effect models were used depending on the heterogeneity between estimates.

**Results:** a total of fifteen studies with 21,614 patients were included. The use of RAS inhibitors was associated with a non-significant 20% decreased risk of the composite outcome (death, admission to intensive care unit, mechanical ventilation requirement or progression to severe or critical pneumonia): RR 0.81 (95%CI: 0.63-1.04), p=0.10, I^2^=82%. In a subgroup analysis that included hypertensive subjects only, ACEI/ARB were associated with a 27% significant decrease in the risk of the composite outcome (RR 0.73 (95%CI: 0.56-0.96), p=0.02, I^2^=65%).

**Conclusion:** the results of this pooled analysis suggest that the use of ACEI/ARB does not worsen the prognosis, and could even be protective in hypertensive subjects. Patients should continue these drugs during their COVID-19 illness.

## 1. Introduction

The coronavirus disease 2019 (COVID-19) outbreak originated in Wuhan in December 2019 and caused by the betacoronavirus SARS-CoV-2, was declared a pandemic by the World Health Organization in March 2020, affecting more than 3,400,000 people and causing more than 240,000 deaths.[1]

Interestingly, COVID-19 seems to manifest as a more severe disease in people with cardiovascular comorbidities, such as hypertension,[2,3] although is not yet very clear whether this association is independent from advanced age.[4] Myocardial injury has been proposed as the link between the inflammatory pathogenesis during the progress of the disease and the poorer prognosis.[5,6] It has been postulated that the virus could damage myocardial cells through several mechanisms including direct damage and systemic inflammatory responses.[6] Subjects with preexisting cardiovascular diseases might be more susceptible to COVID-19–induced heart injury.

Sars-Cov-2 gains entrance to cells through the angiotensin-converting enzyme 2 (ACE2),[7] a carboxypeptidase that converts angiotensin II into angiotensin-(1-7) and counterbalances the renin-angiotensin-aldosterone system, exerting protective effects in the cardiovascular system. Given that there are limited reports that ACE inhibitors affect the expression of ACE2 in the heart and the kidney,[8] there has been a growing concern about angiotensin-converting enzyme inhibitors (ACEI) and angiotensin receptor blockers (ARB) increasing patient susceptibility to viral host cell entry and propagation.[9,10] Of note, many patients with cardiovascular comorbidities are treated with these drug classes. On the other hand, it is hypothesized that SARS-CoV-2, like SARS-CoV, not only gains initial entry through ACE2 but also subsequently downregulates ACE2 expression,[11] and deregulated ACE2 may theoretically mediate acute lung injury.[12] In fact, some experts have advocated for the use of ACEI and ARB to prevent organ injury and there are currently eleven registered clinical trials that will evaluate the potential benefit of ARB or ACEI in either hospitalized or not hospitalized COVID-19 infected patients.

To date, there is insufficient clinical or scientific evidence to recommend the discontinuation or maintenance of ACEI/ARB treatment in face of COVID-19. Therefore, in this article, we conducted a systematic literature search to determine a possible association between the use of ACEI/ARB and the progression of COVID-19 to severe forms or death.

## 2. Methods

Preferred Reporting Items for Systematic Reviews and Meta-Analyses (PRISMA) statement[13] was followed for the conduct and reporting of this systematic review (PRISMA checklist provided as supplementary material).

### 2.1 Data source, search strategy and elegibility criteria

To identify publications regarding the clinical outcomes of COVID-19 in infected patients under treatment or not under treatment with ACEI/ARB, an extensive search of the literature was conducted in Medline (through PubMed interface), Cochrane Library, Google Scholar and the preprint servers for the health sciences MedRxiv and BioRxiv, from December 2019 to May, 2^nd^ 2020. In addition, we manually searched from the reference lists of all relevant retrieved studies (snowball technique) to identify any other studies that may have been missed by our search strategy.

The following search strategy was implemented:

#1: SARS-CoV-2 OR COVID 19 OR coronavirus disease 2019 OR coronavirus 2 OR novel coronavirus OR 2019-nCoV OR receptor to SARS-CoV-2 OR coronavirus entry OR virulence of SARS-CoV-2
#2: hypertension OR blood pressure OR antihypertensive OR angiotensin OR angiotensin converting enzyme inhibitor OR angiotensin receptor blocker OR ACEi OR ARB OR antihypertensive drugs OR withdrawal of RAS inhibitors OR medication use in COVID-19 OR initiation or discontinuation of RAS blockade OR angiotensin-converting enzyme 2 OR ace2 OR renin-angiotensin system OR RAS OR RAS blockers OR RAS treatment OR RAS activity OR cardiovascular disease OR cardiovascular conditions OR diabetes OR chronic kidney disease OR renal disease OR heart failure OR myocardial infarction OR comorbidity
#3: cardiac injury OR respiratory failure OR mortality OR fatality rates OR death OR complications OR survival OR prognosis OR respiratory support OR mechanical ventilation OR noninvasive ventilation OR intensive care unit OR ICU OR cardiomyopathy OR pneumonia OR acute respiratory distress OR ARDS OR myocarditis OR pulmonary disease
#1 AND #2 AND #3

Articles were limited to human studies, original articles including comparative studies, both randomized controlled trials (RCTs) or non-RCTs where treatment with ACEI and/or ARB was compared with no treatment or other antihypertensive drugs in terms of COVID-19 infection serious adverse outcomes. Case reports, non-human studies and studies without adequate information were excluded. Abstracts of articles were then scanned for relevance.

### 2.2 Outcome measures

Our outcome was a combination of death, admission to intensive care unit (ICU), mechanical ventilation requirement or progression to severe or critical pneumonia. According to the Report of the WHO-China Joint Mission on Coronavirus Disease 2019 (COVID-19),[14] severe pneumonia was defined as dyspnea, respiratory frequency ≥30/minute, blood oxygen saturation ≤93%, PaO2/FiO2 ratio <300, and/or lung infiltrates >50% of the lung field within 24-48 hours whereas critical pneumonia was defined as respiratory failure, septic shock, and/or multiple organ dysfunction/failure and critical pneumonia as respiratory failure, septic shock, and/or multiple organ dysfunction/failure.

### 2.3 Data extraction and study quality assessment

Data were collected by two independent reviewers (RM and JB) and entered into a predesigned data extraction form. Differences between the two reviewers regarding study eligibility were resolved by consensus.

Study and population characteristics were extracted from each included study. Regarding study characteristics, the name of the first author, the date of publication, the study type, the number of centers and countries contributing to the research as well as possible funding were extracted. Also, total population, mean age, percentage of male participants, number of patients taking ACEI and/or ARB, number of patients taking other antihypertensives or no treatment, type and dose of antihypertensive drug, number of patients experiencing the primary outcome in each treatment group as well as details for possible matched characteristics were recorded. Finally, the presence of comorbidities such as hypertension, diabetes, cardio and cerebrovascular disease were also extracted.

The Newcastle-Ottawa Scale (NOS) was used to assess the quality of studies.[15] In this scale, a ‘star system’ is used, in which a study is judged on three aspects: selection of the study groups; comparability of the groups; and ascertainment of either the exposure or outcome of interest for case-control or cohort studies, respectively. A study can be awarded a maximum of one star for each numbered item within the selection and exposure categories. A maximum of two stars can be given for comparability. In this systematic review, studies with scores above 6 were considered as high quality, 3-6 as moderate and those with scores below than 3 as low quality.[16]

### 2.4 Data synthesis and statistical analysis

Pooled analysis was performed to estimate the risk ratio (RR) and 95% confidence interval (95% CI) comparing the occurrence of the composite outcome in COVID-19 infected patients under treatment with ACEI/ARB vs no treatment or treatment other than ACEI/ARB. All analyses were conducted using RevMan software version 5.3. Fixed-effect models or random-effect models were used depending on the heterogeneity between estimates. We used the I^2^ statistics to assess the magnitude of heterogeneity. The fixed effect model was used if I^2^<50% and the random effect model was used if I^2^≥50%.

Publication bias was examined by the use of a funnel plot of each study’s effect size against the precision (1/SE).

We planned to conduct three subgroup analyses: a) including only high quality studies; b) including only peer-reviewed published studies; c) including only hypertensive subjects.

## 3. Results

### 3.1 Search Results and Study Characteristics

Our comprehensive search initially identified 798 articles. The flowchart diagram in Figure 1 shows our literature search, study selection, and the number of studies included. After removing duplicates, reviews, letters, guidelines or consensus, and studies not addressing the research question, 24 full-text articles were assessed for eligibility. Among them, 17 studies that satisfied the eligibility criteria were included in this systematic review. After unsuccessfully trying to contact the authors, we had to exclude two of these studies from the quantitative analysis because the data of interest were not extractable. Therefore, 15 studies were finally included in the meta-analysis, totaling 21,614 COVID-19 patients. Among them, 6477 were taking ACEI or ARB and 15,137 were under other or no treatment.

**Fig. 1.**
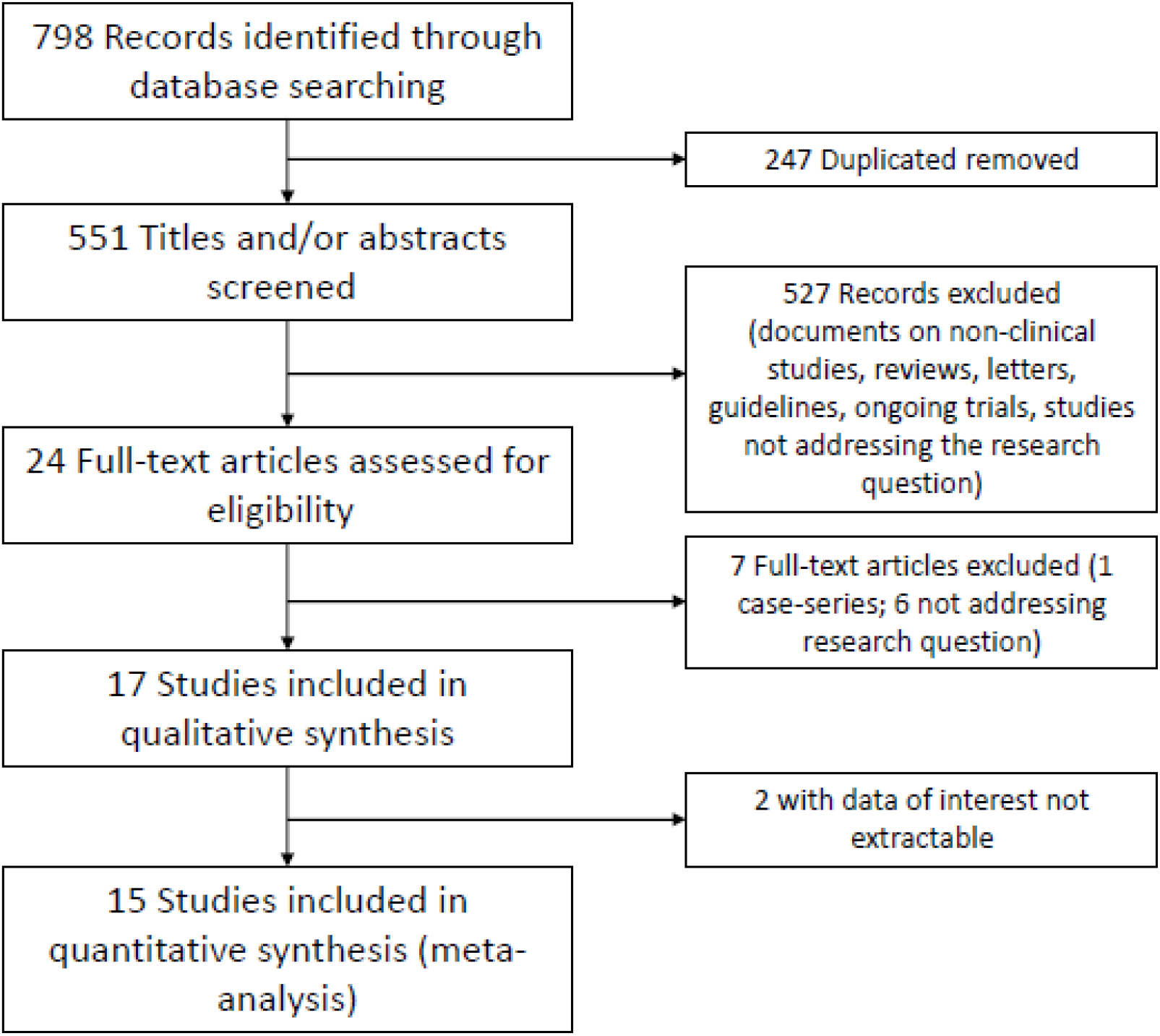
Flowchart depicting literature review and study selection.

All of the selected studies were published in 2020 and were all of observational nature.[17*33] In nine studies, the outcome that matched our research question was death;[20,23,25*28,31*33] in two studies, severe pneumonia;[19,21] in one study, critical or severe pneumonia;[18] in one study, severe pneumonia or death;[22] in one study, critical pneumonia or death [29]; in one study, admission to ICU;[24] in one study, death or admission to ICU [17] and in one study, death, admission to ICU or mechanical ventilation requirement.[30] In three of the studies where the outcomes death and severe pneumonia,[26,27] or critical pneumonia [25] were discriminated, we chose mortality for our analysis since the data necessary to evaluate the outcome as a composite were not available. Table 1 summarizes the study characteristics. Quality assessment through the Newcastle-Ottawa Scale showed scores above 6 points (between 7 and 9), indicating high quality studies, except for one study that scored 6 points (moderate quality).

**Table 1.**
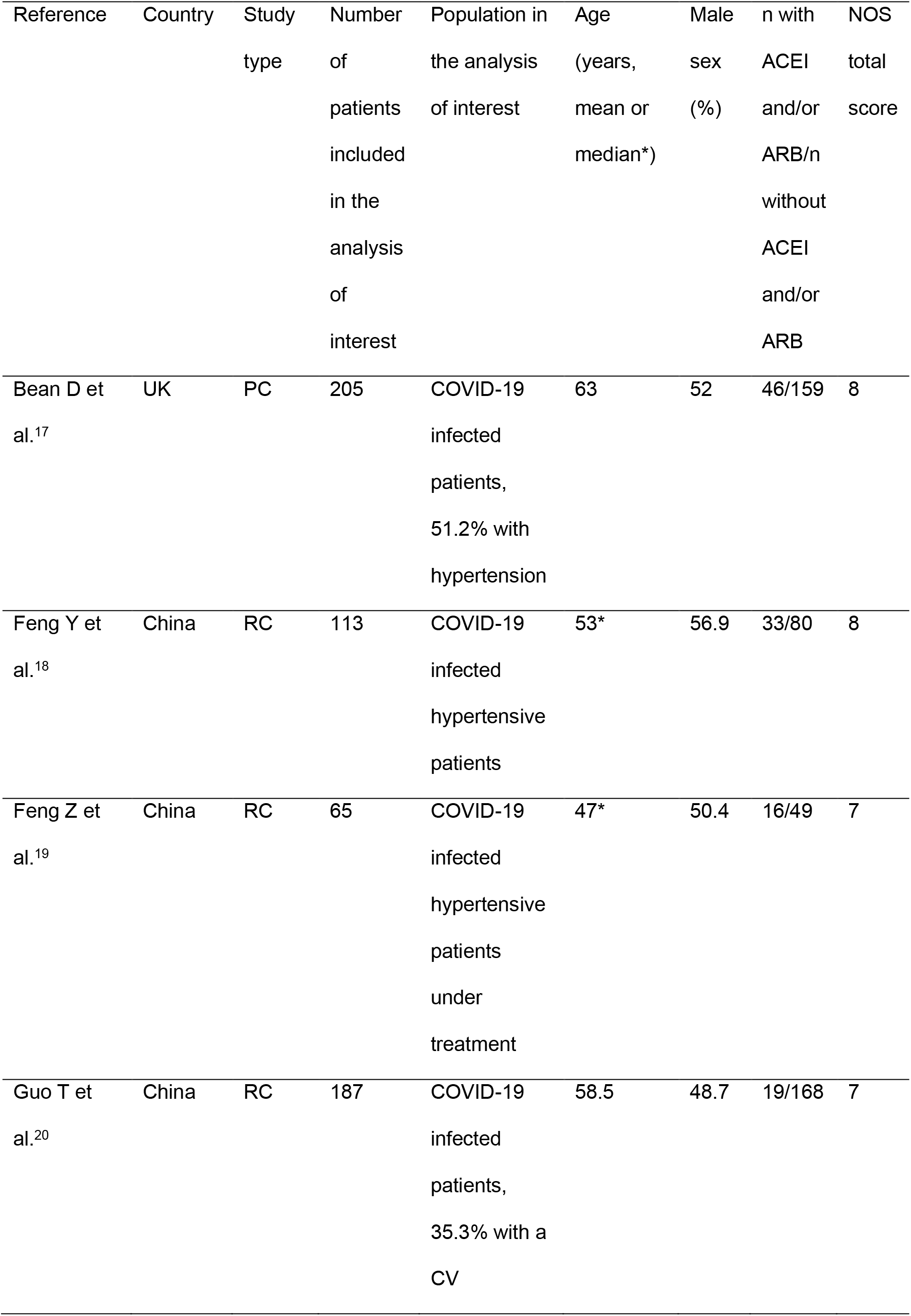
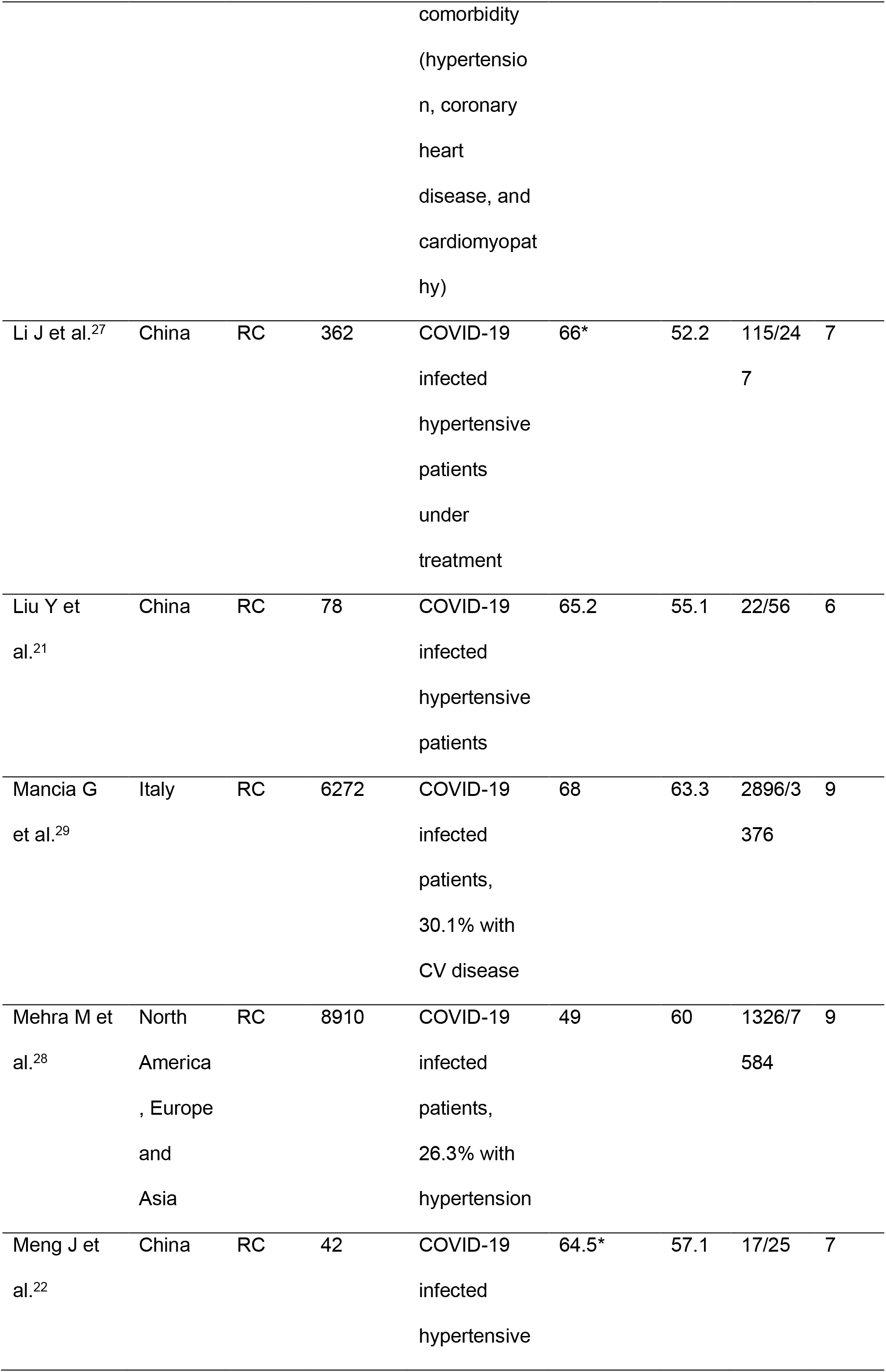
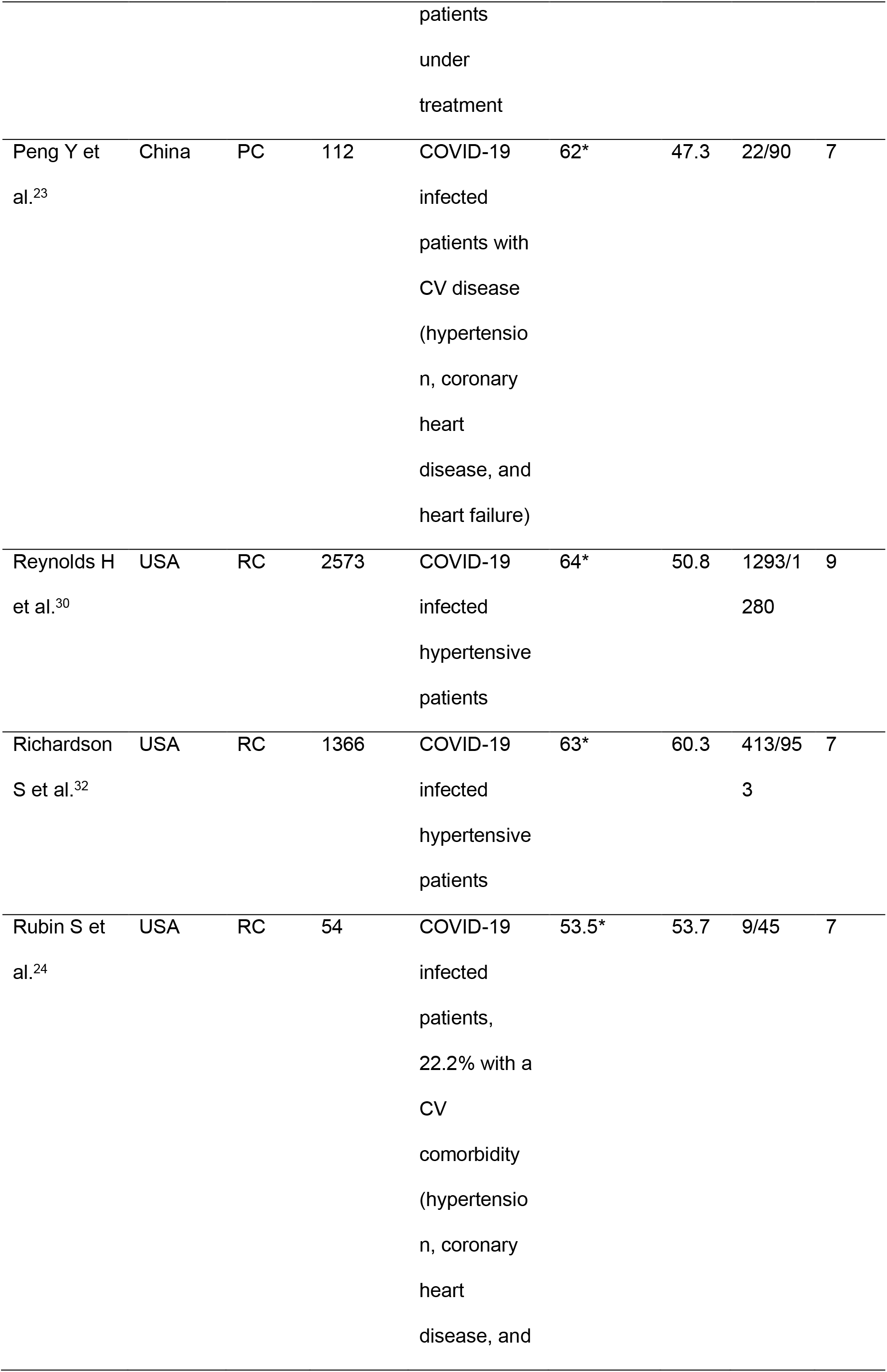
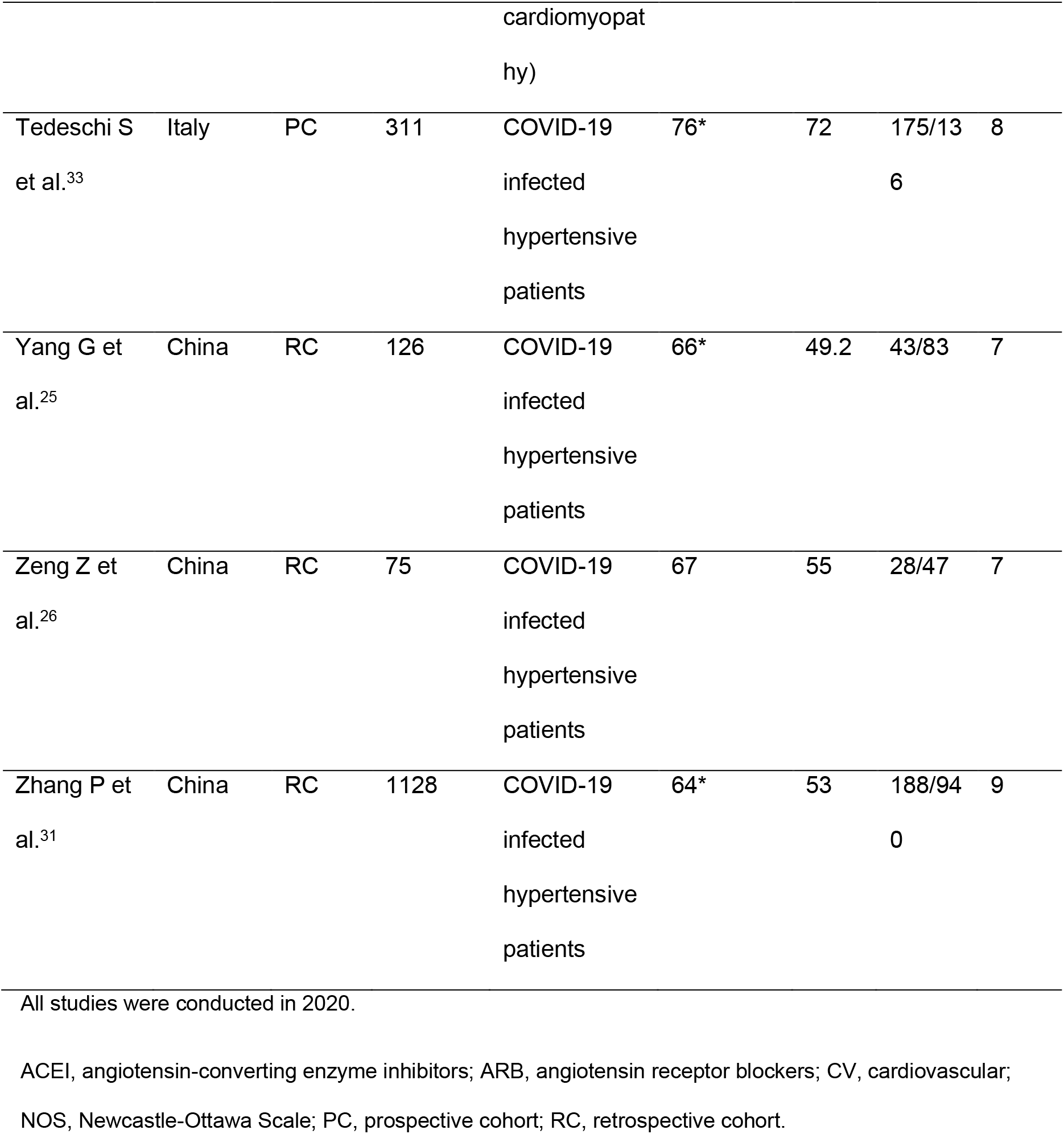
Description of the included studies.

In a qualitative synthesis, one study showed a higher risk of the outcome with the use of ACEI/ARB,[29] three studies showed a lower risk,[18,28,31] and thirteen studies found no association.[17,19*27,30,32,33]

### 3.2 Meta-analysis

Among the 17 studies included in the systematic review, we had to exclude two studies [24,33] from the meta-analysis because the data of interest were not extractable. The results of our pooled analysis for the identified studies are presented in Figure 2. The use of ACEI or ARB was found to decrease the risk of the composite outcome (death, admission to ICU, mechanical ventilation requirement or progression to severe or critical pneumonia) in nearly 20%, although this difference did not reach statistical significance: RR 0.81 (95%CI: 0.63-1.04), test for overall effect: Z=1.66, p=0.10. High heterogeneity was observed in the analysis: I^2^=82%, Chi^2^=77.46, df=14, p<0.001).

**Fig. 2.**
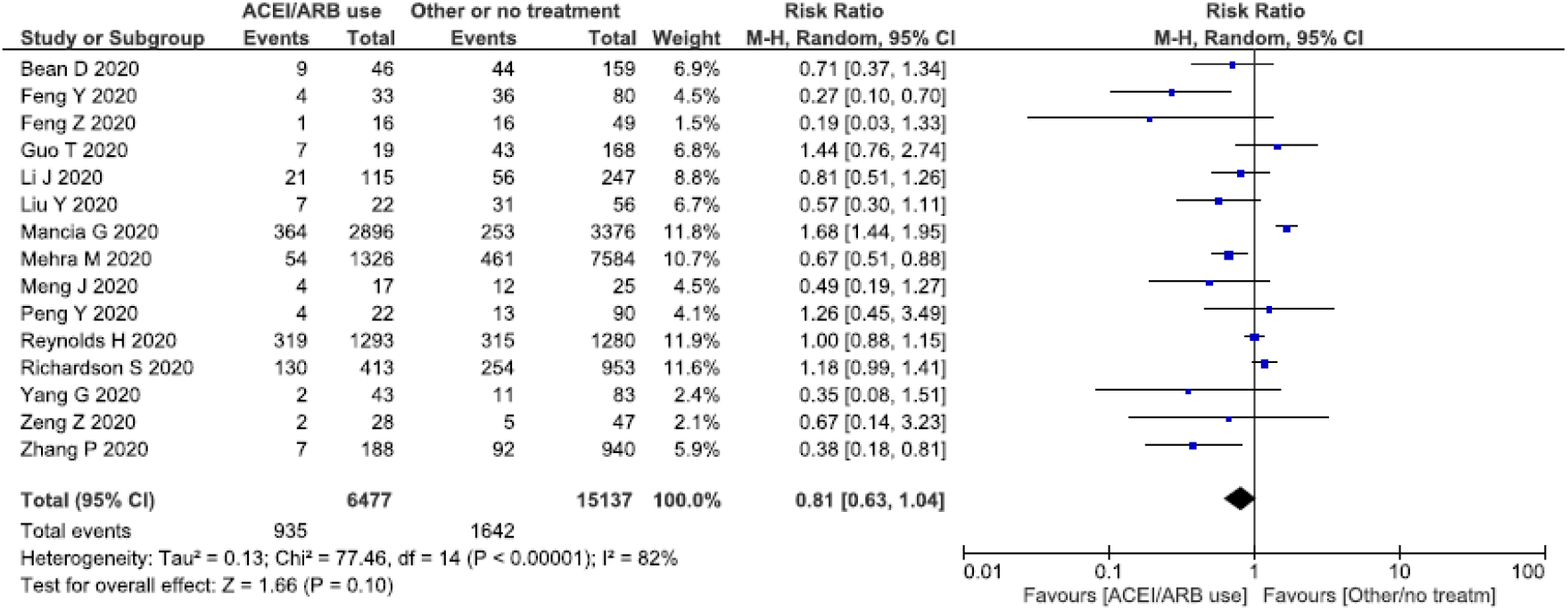
Forest plot showing risk ratios (RRs) of the composite outcome (death, admission to intensive care unit (ICU), mechanical ventilation requirement or progression to severe or critical pneumonia)

In our subgroup analyses, no significant differences were found when excluding the study with a NOS score below 7 or when excluding preprints. Of note, in the subgroup analysis that included only hypertensive subjects, the use of ACEI/ARB was found to significantly reduce the risk of the composite outcome: RR 0.73 (95%CI: 0.56-0.96), test for overall effect: Z=2.25, p=0.02; I^2^=65%, Chi2=28.69, df=10, p=0.001). (Figure 3).

**Fig. 3.**
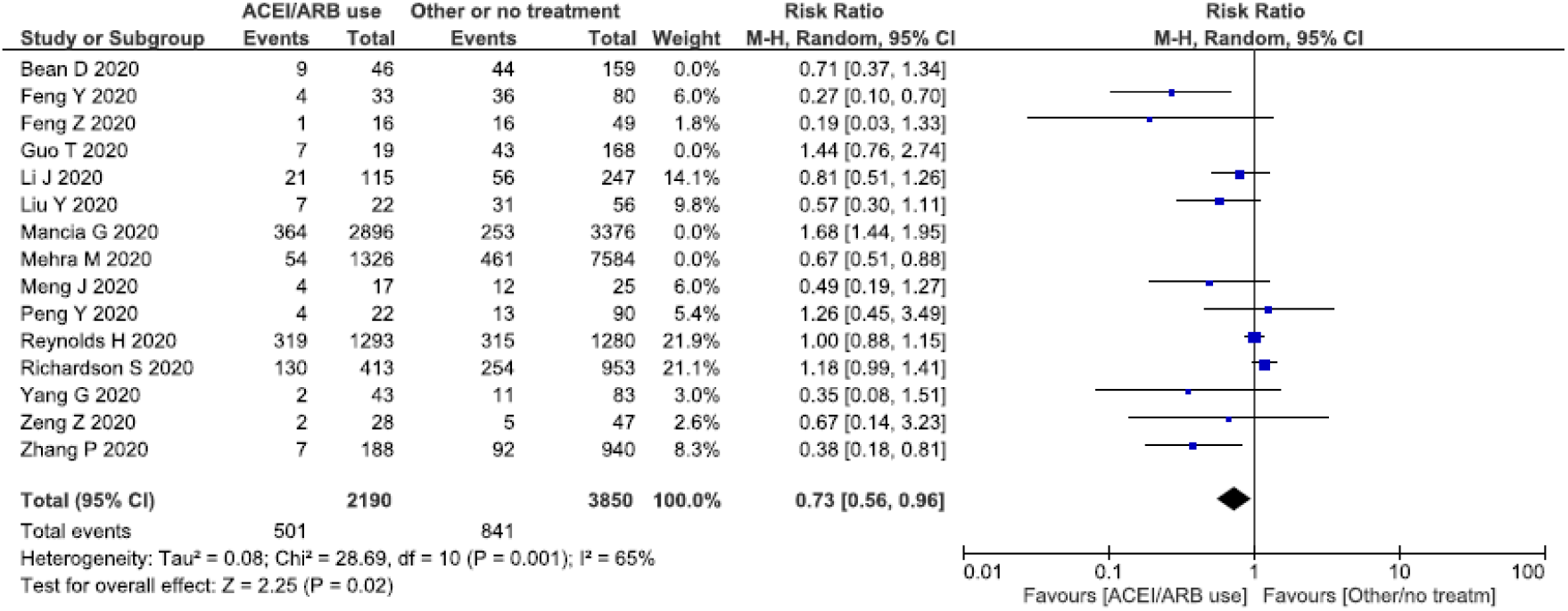
Sensitivity analysis including studies on hypertensive subjects only.

Finally, the funnel plot indicated some degree of publication bias, with a lack of small studies showing increased risk of ACEI/ARB use (Figure S1).

## 4. Discussion

In this meta-analysis we found that the use of ACEI/ARB was not associated with an increased risk of death, admission to intensive care unit (ICU), mechanical ventilation requirement or progression to severe or critical pneumonia in COVID-19 infected patients. In fact, we observed a tendency (although non-significant) towards a protective effect.

Controversy has arisen in the scientific community regarding the use of RAS inhibitors in the context of COVID-19: some experts hypothesized that these drugs might increase susceptibility to the virus by increasing the expression of ACE2, the SARS-CoV2 receptor for host cell entry.[9,10] On the other hand, scientific societies recommend not to discontinue RAS inhibitors because of insufficient evidence of their potential harm and overwelming evidence on their benefits [34*38] and even some researchers advocate for the use of RAS inhibitors based on the fact that, through increasing the expression of ACE2, they counterbalance the reduction in pulmonary ACE2 provoked by the virus either through internalization with viral entry and/or downregulation of ACE2 enzyme during this process, which would lower the production of inflammatory cytokines,[39] thus exerting a protective role in lung injury.[40] Remarkably, RAS inhibitors have been shown to be associated with reduced mortality in patients with sepsis.[41] A study conducted by Henry C et al. has found a beneficial effect of ACEI/ARB in patients admitted with viral pneumonia, as they significantly reduced the pulmonary inflammatory response and cytokine release caused by virus infection.[42] Moreover, other studies have shown that these drugs would attenuate the inflammatory response in COVID-19 infected patients, possibly through the inhibition of IL-6 levels,[22] C-reactive protein and procalcitonin.[25]

Recent evidence has consistently shown that hypertension and other cardiovascular morbidities are more frequent in COVID-19 infected patients who carry the worst prognosis.[2,3,43] Of note, most of these studies did not make adjustments for important co-variates such as age, although more recent studies are starting to show that this association holds even after such adjustments.[44,45] As many subjects with cardiovascular morbidities are under treatment with ACEI/ARB, it has been speculated that the use of these drugs could be the underlying cause of the relation between these morbidities and a poorer COVID-19 prognosis. If this was the case, we would have found a greater risk of severe forms of COVID-19 in subjects under treatment with ACEI/ARB, but we have not. In fact, the evidence presented in this meta-analysis supports the idea that ACEI/ARB should not be discontinued when treating COVID-19 patients with cardiovascular morbidities.

On the other hand, RAS inhibitors have established benefits in protecting the cardiovascular system.[46,47] Their discontinuation may increase the chance of decompensation in high-risk patients as the benefits that are specific to these drugs may not be offset by other antihypertensive agents. For instance, it has been shown that withdrawal of ACEI/ARB during heart failure hospitalization is associated with higher rates of postdischarge mortality.[48] Besides, antihypertensive medication changes would require frequent dose adjustment and management of adverse effects, increasing the need for patients to visit their doctors and, therefore, the exposure to COVID-19 and risk of infection.

Clearly, more research is needed to elucidate the multifacetic role of the RAS in connection with SARS-CoV-2 infection. The evidence regarding the use of RAS inhibitors in patients with COVID-19 infection is still emerging. Currently, in patients who previously used ACEI/ARB, the use of these drugs may not need to be discontinued in order to prevent COVID-19 complications.

Finally, our findings must be interpreted in the context of the meta-analysis limitations. First, we could not make a distinction between the effects of ACEI and ARB separately, since most studies evaluated the two drug classes together. Second, all the studies included in the meta-analysis were observational and some were preprints. We could not find any randomized clinical trials that already showed results addressing our research question. On the other hand, in planning future trials, randomizing subjects to discontinue drugs with proven benefits would probably raise ethical concerns. Third, most of the included observational studies were retrospective cohorts and potential selection bias of patients is an indisputable concern. Fourth, residual confounders may be present in these observational studies and, even when measured, many studies did not make adjustments for such potential confounders. It must be noted, however, that not adjusting for age and cardiovascular morbidities would shift the results towards a higher risk since these variables are positively associated with the use of ACEI/ARB and with COVID-19 poorer outcomes.

In conclusion, large prospective studies are required to confirm our finding and to explore the mechanisms for a possible protective role of RAS inhibitors in the context of COVID-19. In the meantime, these early results suggest that patients on ACEI/ARB should continue their treatment during COVID-19 illness, supporting current recommendations from multiple scientific societies.

## Data Availability

The datasets generated during and/or analysed during the current study are available from the corresponding author on reasonable request.

https://www.researchgate.net/publication/341489408_Data_search_strategy

## Ethics approval

this is a systematic review and meta-analysis of the literature. No human subjects or animals were involved in this research.

## List of Supplemental Digital Content

**-Fig. S1.**
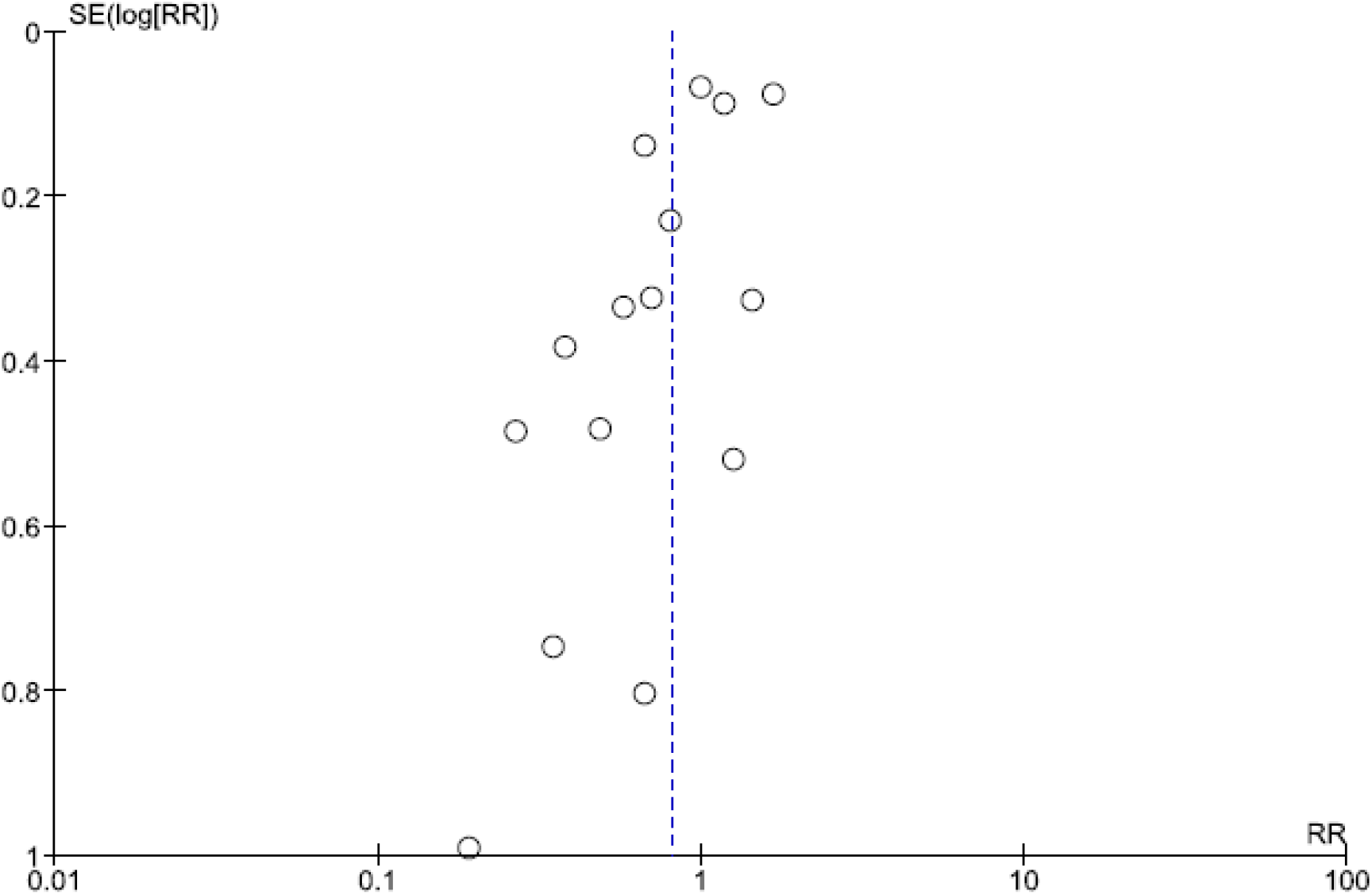
Funnel plot of the comparison of the composite outcome between patients under ACEI/ARB and not under ACEI/ARB

## References

1. https://gisanddata.maps.arcgis.com/apps/opsdashboard/index.html#/bda7594740fd40299423467b48e9ecf6. Last accessed: 3/5/2020.

2. Wu C, Chen X, Cai Y, et al. Risk factors associated with acute respiratory distress syndrome and death in patients with coronavirus disease 2019 pneumonia in Wuhan, China. JAMA Intern Med. Published online March 13, 2020.

3. Zhou F, Yu T, Du R, et al. Clinical course and risk factors for mortality of adult inpatients with COVID-19 in Wuhan, China: a retrospective cohort study. Lancet. 2020;S0140–6736(20)30566-3.

4. Sommerstein R, Kochen MM, Messerli FH, Gräni C. Coronavirus Disease 2019 (COVID-19): Do Angiotensin-Converting Enzyme Inhibitors/Angiotensin Receptor Blockers Have a Biphasic Effect? J Am Heart Assoc. 2020 Apr 7;9(7):e016509.

5. Shi S, Qin M, Shen B, Cai Y, Liu T, Yang F, et al. Association of Cardiac Injury With Mortality in Hospitalized Patients With COVID-19 in Wuhan, China. JAMA Cardiol. 2020 Mar 25.

6. Huang C, Wang Y, Li X, Ren L, Zhao J, Hu Y, et al. Clinical features of patients infected with 2019 novel coronavirus in Wuhan, China. Lancet. 2020 Feb 15;395(10223):497–506.

7. Letko M, Marzi A, Munster V. Functional assessment of cell entry and receptor usage for SARS-CoV-2 and other lineage B betacoronaviruses. Nat Microbiol. 2020 Apr;5(4):562–569.

8. Ferrario CM, Jessup J, Chappell MC, Averill DB, Brosnihan KB, Tallant EA, et al. Effect of angiotensin-converting enzyme inhibition and angiotensin II receptor blockers on cardiac angiotensin-converting enzyme 2. Circulation. 2005 May 24;111 (20):2605–10.

9. Esler M, Esler D. Can angiotensin receptor-blocking drugs perhaps be harmful in the COVID-19 pandemic? J Hypertens. 2020 May;38(5):781–782.

10. Diaz JH. Hypothesis: angiotensin-converting enzyme inhibitors and angiotensin receptor blockers may increase the risk of severe COVID-19. J Travel Med. 2020 Mar 18. pii: taaa041.

11. Kuba K, Imai Y, Rao S, Gao H, Guo F, Guan B, et al. A crucial role of angiotensin converting enzyme 2 (ACE2) in SARS coronavirus-induced lung injury. Nat Med. 2005 Aug;11(8):875–9.

12. Liu Y, Yang Y, Zhang C, Huang F, Wang F, Yuan J, et al. Clinical and biochemical indexes from 2019-nCoV infected patients linked to viral loads and lung injury. Sci China Life Sci. 2020 Mar;63(3):364–374.

13. Moher D, Liberati A, Tetzlaff J, Altman DG; PRISMA Group. Preferred reporting items for systematic reviews and meta-analyses: the PRISMA statement. J Clin Epidemiol. 2009 Oct;62(10):1006–12.

14. https://www.who.int/docs/default-source/coronaviruse/who-china-joint-mission-on-covid-19-final-report.pdf. Last accessed: 14/4/2020.

15. Wells G, Shea B, O’connell D, Peterson J, Welch V, Losos M, et al. The Newcastle-Ottawa Scale (NOS) for assessing the quality of nonrandomised studies in meta-analyses. http://www.ohri.ca/programs/clinical_epidemiology/oxford.asp. Last accessed: 14/4/2020.

16. Behboudi-Gandevani S, Amiri M, Bidhendi Yarandi R, Ramezani Tehrani F. The impact of diagnostic criteria for gestational diabetes on its prevalence: a systematic review and meta-analysis. Diabetol Metab Syndr. 2019 Feb 1;11:11.

17. Daniel Bean, Zeljko Kraljevic, Thomas Searle, Rebecca Bendayan, Andrew Pickles, Amos Folarin, et al. Treatment with ACE-inhibitors is associated with less severe disease with SARS-Covid-19 infection in a multi-site UK acute Hospital Trust. medRxiv 2020.04.07.20056788; doi: https://doi.org/10.1101/2020.04.07.20056788 (preprint).

18. Feng Y, Ling Y, Bai T, Xie Y, Huang J, Li J, et al. COVID-19 with Different Severity: A Multi-center Study of Clinical Features. Am J Respir Crit Care Med. 2020 Apr 10. doi: 10.1164/rccm.202002-0445OC.

19. Zhichao Feng, Jennifer Li, Shanhu Yao, Qizhi Yu, Wenming Zhou, Xiaowen Mao, et al. The Use of Adjuvant Therapy in Preventing Progression to Severe Pneumonia in Patients with Coronavirus Disease 2019: A Multicenter Data Analysis. medRxiv 2020.04.08.20057539; doi: https://doi.org/10.1101/2020.04.08.20057539 (preprint).

20. Guo T, Fan Y, Chen M, Wu X, Zhang L, He T, et al. Cardiovascular Implications of Fatal Outcomes of Patients With Coronavirus Disease 2019 (COVID-19). JAMA Cardiol. 2020 Mar 27.

21. Yingxia Liu, Fengming Huang, Jun Xu, Penghui Yang, Yuhao Qin, Mengli Cao, et al. Anti-hypertensive Angiotensin II receptor blockers associated to mitigation of disease severity in elderly COVID-19 patients. medRxiv 2020.03.20.20039586; doi: https://doi.org/10.1101/2020.03.20.20039586 (preprint).

22. Meng J, Xiao G, Zhang J, He X, Ou M, Bi J, et al. Renin-angiotensin system inhibitors improve the clinical outcomes of COVID-19 patients with hypertension. Emerg Microbes Infect. 2020 Dec;9(1):757–760.

23. Peng YD, Meng K, Guan HQ, Leng L, Zhu RR, Wang BY, et al. [Clinical characteristics and outcomes of 112 cardiovascular disease patients infected by 2019-nCoV]. Zhonghua Xin Xue Guan Bing Za Zhi. 2020 Mar 2;48(0):E004.

24. Samuel J. S. Rubin, Samuel Robert Falkson, Nicholas Degner, Catherine Blish. Clinical characteristics associated with COVID-19 severity in California. medRxiv 2020.03.27.20043661; doi:https://doi.org/10.1101/2020.03.27.20043661 (preprint).

25. Yang G, Tan Z, Zhou L, Yang M, Peng L, Liu J, Cai J, Yang R, Han J, Huang Y, He S. Effects Of ARBs And ACEIs On Virus Infection, Inflammatory Status And Clinical Outcomes In COVID-19 Patients With Hypertension: A Single Center Retrospective Study. Hypertension. 2020 Apr 29. doi:10.1161/HYPERTENSIONAHA.120.15143..

26. Zhenhua Zeng, Tong Sha, Yuan Zhang, Feng Wu, Hongbin Hu, Haijun Li, et al. Hypertension in patients hospitalized with COVID-19 in Wuhan, China: A single-center retrospective observational study. medRxiv 2020.04.06.20054825; doi: https://doi.org/10.1101/2020.04.06.20054825 (preprint).

27. Li J, Wang X, Chen J, Zhang H, Deng A. Association of Renin-Angiotensin System Inhibitors With Severity or Risk of Death in Patients With Hypertension Hospitalized for Coronavirus Disease 2019 (COVID-19) Infection in Wuhan, China. JAMA Cardiol. 2020 Apr 23.

28. Mehra MR, Desai SS, Kuy S, Henry TD, Patel AN. Cardiovascular Disease, Drug Therapy, and Mortality in Covid-19. N Engl J Med. 2020 May 1. doi:10.1056/NEJMoa2007621. [Epub ahead of print]

29. Mancia G, Rea F, Ludergnani M, Apolone G, Corrao G. Renin-Angiotensin-Aldosterone System Blockers and the Risk of Covid-19. N Engl J Med. 2020 May 1. doi: 10.1056/NEJMoa2006923. [Epub ahead of print]

30. Reynolds HR, Adhikari S, Pulgarin C, Troxel AB, Iturrate E, Johnson SB, et al. Renin-Angiotensin-Aldosterone System Inhibitors and Risk of Covid-19. N Engl J Med. 2020 May 1. doi: 10.1056/NEJMoa2008975. [Epub ahead of print]

31. Zhang P, Zhu L, Cai J, Lei F, Qin JJ, Xie J, et al. Association of Inpatient Use of Angiotensin Converting Enzyme Inhibitors and Angiotensin II Receptor Blockers with Mortality Among Patients With Hypertension Hospitalized With COVID-19. Circ Res. 2020 Apr 17.doi: 10.1161/CIRCRESAHA.120.317134. [Epub ahead of print]

32. Richardson S, Hirsch JS, Narasimhan M, Crawford JM, McGinn T, Davidson KW, et al. Presenting Characteristics, Comorbidities, and Outcomes Among 5700 Patients Hospitalized With COVID-19 in the New York City Area. JAMA. 2020 Apr 22. doi:10.1001/jama.2020.6775. [Epub ahead of print]

33. Tedeschi S, Giannella M, Bartoletti M, Trapani F, Tadolini M, Borghi C, et al. Clinical impact of renin-angiotensin system inhibitors on in-hospital mortality of patients with hypertension hospitalized for COVID-19. Clin Infect Dis. 2020 Apr 27. pii: ciaa492. doi: 10.1093/cid/ciaa492. [Epub ahead of print]

34. https://www.eshonline.org/spotlights/esh-statement-covid-19/. Last accessed: 17/4/20.

35. https://www.escardio.org/Councils/Council-on-Hypertension-(CHT)/News/position-statement-of-the-esc-council-on-hypertension-on-ace-inhibitors-and-ang. Last accessed: 17/4/20.

36. https://www.ccs.ca/images/Images2020/CCSCHFSstatementregardingCOVIDEN.pdf. Last accessed: 17/4/20.

37. https://ish-world.com/news/a/A-statement-from-the-International-Society-of-Hypertension-on-COVID-19/. Last accessed: 17/4/20.

38. https://professional.heart.org/professional/ScienceNews/UCM505836HFSAACCAHAstatement-addresses-concerns-re-using-RAAS-antagonists-in-COVID.jsp. Last accessed: 17/4/20.

39. Ferrario CM, Strawn WB. Role of the renin-angiotensin-aldosterone system and proinflammatory mediators in cardiovascular disease. Am J Cardiol. 2006 Jul 1; 98(1):121–8.

40. Huang F, Guo J, Zou Z, Liu J, Cao B, Zhang S, et al. Angiotensin II plasma levels are linked to disease severity and predict fatal outcomes in H7N9-infected patients. Nat Commun. 2014 May 6;5:3595.

41. Hsu WT, Galm BP, Schrank G, Hsu TC, Lee SH, Park JY, et al. Effect of Renin-Angiotensin-Aldosterone System Inhibitors on Short-Term Mortality After Sepsis: A Population-Based Cohort Study. Hypertension. 2020 Feb;75(2):483–491.

42. Henry C, Zaizafoun M, Stock E, Ghamande S, Arroliga AC, White HD. Impact of angiotensin-converting enzyme inhibitors and statins on viral pneumonia. Proc (Bayl Univ Med Cent). 2018 Oct 26;31 (4):419–423.

43. Guan WJ, Ni ZY, Hu Y, Liang WH, Ou CQ, He JX, et al; China Medical Treatment Expert Group for Covid-19. Clinical Characteristics of Coronavirus Disease 2019 in China. N Engl J Med. 2020 Feb 28.

44. Wang L, He W, Yu X, Hu D, Bao M, Liu H, et al. Coronavirus disease 2019 in elderly patients: Characteristics and prognostic factors based on 4-week follow-up. J Infect. 2020 Mar 30. pii: S0163–4453(20)30146-8.

45. Guan WJ, Liang WH, Zhao Y, Liang HR, Chen ZS, Li YM, et al; China Medical Treatment Expert Group for Covid-19. Comorbidity and its impact on 1590 patients with Covid-19 in China: A Nationwide Analysis. Eur Respir J. 2020 Mar 26. pii:2000547.

46. Vaduganathan M, Vardeny O, Michel T, McMurray JJV, Pfeffer MA, Solomon SD. Renin-Angiotensin-Aldosterone System Inhibitors in Patients with Covid-19. N Engl J Med. 2020 Mar 30.

47. Danser AHJ, Epstein M, Batlle D. Renin-Angiotensin System Blockers and the COVID-19 Pandemic: At Present There Is No Evidence to Abandon Renin-Angiotensin System Blockers. Hypertension. 2020 Mar 25:HYPERTENSIONAHA12015082.

48. Gilstrap LG, Fonarow GC, Desai AS, Liang L, Matsouaka R, DeVore AD, et al. Initiation, Continuation, or Withdrawal of Angiotensin-Converting Enzyme Inhibitors/Angiotensin Receptor Blockers and Outcomes in Patients Hospitalized With Heart Failure With Reduced Ejection Fraction. J Am Heart Assoc. 2017 Feb 11; 6(2). pii: e004675.

